# Implementation of Pre-emptive Pharmacogenomics Testing in Outpatient Clinics in Asia (IMPT study)

**DOI:** 10.1101/2024.07.19.24310681

**Authors:** Fiona FJ Ng, Rashmi Verma, Levana Sani, Astrid Irwanto, Michael Lee, Angeline Wee, Shih Kiat Chng, Melvyn Wong, Alexandre Chan

**Author notes:** Corresponding Author (AC).

## Abstract

**Background:** In view of the limited data related to preemptive pharmacogenomics (PGx) testing in the primary care setting, we designed a study to assess the feasibility of implementing preemptive PGx services at outpatient clinics, with the aim to assess the practicality and challenges of implementing preemptive PGx testing within primary care, and its impact on clinical workflows and patient care.

**Methods:** This prospective study was conducted between October 2022 and August 2023 at five outpatient clinics located in Singapore. Patients aged 21 to 65 with a reported history or risk of developing any of the target chronic conditions or any patients receiving one of the 29 PGx-associated medications were recruited. Patients’ buccal samples were processed using the Nala RxReady™, a multi-gene qPCR-based panel of 21 allele variants of five pharmacogenes. Surveys were administered to study participants and clinicians to assess their perceptions and outcomes related to PGx testing.

**Results:** Among the 222 patients, 95% had at least one clinically actionable variant. Of these patients, 113 reported taking at least one of the 29 studied drugs, with 21.2% of them receiving at least one clinically actionable recommendation based on their PGx results. A total of 150 patients (67.6%) participated in the post-test follow-up survey. Among them, 70% expressed feeling relieved and happy upon receiving their test reports and reported increased confidence in taking their prescribed medication. Furthermore, clinicians identified the necessity for clearer legal regulations regarding PGx testing and insurance coverage to enhance future adoption of PGx testing.

**Conclusions:** Given a high prevalence of clinically actionable variants in almost all tested patients, this study underscores the feasibility and clinical benefits of preemptive PGx testing in primary care clinics.

Clinical Trial Registration: This study is registered with ClinicalTrials.gov, identifier NCT05504135, with the registration date of August 17, 2022.

## Introduction

Pharmacogenomics (PGx) is the study of how an individual’s genetic makeup influences their response to medications. One approach, which is preemptive PGx testing using multi-gene panels, has demonstrated clinical benefits in several studies, primarily conducted in the United States (US) and Europe (Dunnenberger et al., 2015; Gottesman et al., 2013; Ji et al., 2016; Swen et al., 2023; Tsermpini et al., 2020; Wang et al., 2022). In a study conducted among seven European countries encompassing both primary and tertiary care settings, over 90% genotyped patients had at least one actionable PGx variant, and preemptive PGx reduced clinically relevant adverse drug reactions by 30% (Swen et al., 2023). Clearly, preemptive PGx testing and implementation have major impacts on health outcomes that would warrant further evaluation and optimization.

With over 60% of patients visiting primary care in both the UK and the US receiving medications associated with PGx recommendations (Kimpton et al., 2019; Rollinson et al., 2020; Schildcrout et al., 2012), it is vital to fine tune the implementation of preemptive PGx in primary care. One study conducted in the Netherlands revealed that 1 in 19 new prescriptions in the primary care settings would require a dose adjustment or a change in drug therapy (Bank et al., 2019), underscoring the potential utility of preemptive PGx testing in delivering more efficient and effective patient care.

The implementation of PGx into the primary care setting is a multifaceted process that encompasses preemptive PGx research, the integration of PGx into clinical workflows, PGx testing, assessing clinical impact and providing PGx education (Giri et al., 2019). In view of the limited data related to preemptive PGx testing in the outpatient setting, we designed a study to assess the feasibility of implementing preemptive PGx services at outpatient clinics, encompassing both primary and secondary care settings in Singapore. We aimed to explore the practicality and potential challenges of integrating PGx testing, with a particular focus on understanding how the implementation of PGx might impact clinical workflows and patient care.

## Methods

### Study Design

This was a prospective study conducted between October 3, 2022 and August 31, 2023 in three General Practitioner Clinics (Raffles Executive Medical Centre, Raffles Medical at Marina Bay Financial Centre, and Raffles Medical at Singapore Land Tower) and two Specialist Outpatient Clinics (Raffles Diabetes and Endocrine Centre, and Raffles Neuroscience Centre) at Raffles Medical Group (RMG), Singapore. RMG is one of the largest private healthcare enterprises in Singapore, operating a network that spans from primary care at its Raffles Medical clinics to specialist and tertiary care at Raffles Hospital. In 2022, RMG in Singapore provided healthcare services to approximately 40% of the population, reflecting its significant role in the nation’s healthcare landscape (Raffles Medical Group, 2023).

This study was approved by Raffles Hospital Institutional Review Board (Reference number 2017/007). Written informed consent was obtained from all participants and patients in this study.

### Intervention

In this study, we aimed to implement preemptive PGx at outpatient clinics within RMG. Our implementation of PGx testing involved a comprehensive strategy to ensure its successful integration into clinical practice. Our intervention encompassed training, patient recruitment, sample processing, and post-test reporting (Figure 1).

**Figure 1:**
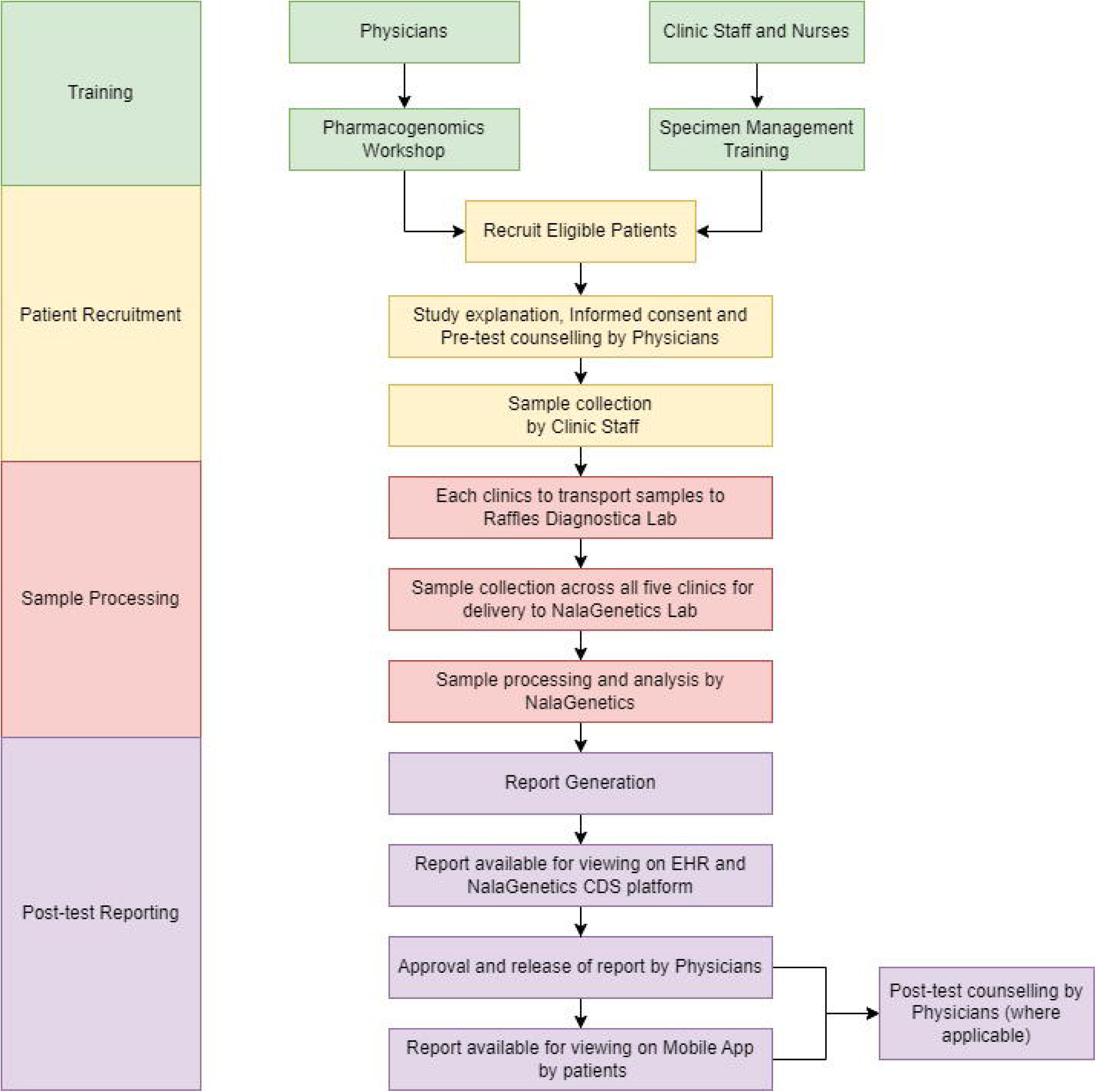
**Integrated Workflow: Training, Patient Recruitment, Sample Processing, and Reporting**

#### a) Training

All participating clinicians (physicians and pharmacists) were required to attend a Pharmacogenomic workshop conducted by Nalagenetics’ Medical Affairs team, which is a comprehensive training program which covers various aspects related to the implementation of preemptive PGX, which includes the significance of PGx testing, the testing process, and PGx results interpretation (Adesta et al., 2021). Following the workshop, participating clinicians were expected to complete a post-training assessment to evaluate their use of PGx in clinical practice. Additionally, all clinic staff underwent specimen management training conducted by Nalagenetics’ Operations team, which included training on sample collection and operational workflow.

#### b) Patient Recruitment

The eligibility criteria included patients aged 21 to 65 with a reported history or risk of developing any of the target chronic conditions: diabetes mellitus, hypertension, dyslipidemia, ischemic heart disease, stroke, osteoarthritis, rheumatoid arthritis, gout, anxiety and major depressive disorder, or any patients who were prescribed with one of the 29 PGx-associated medications (Table 1). Patients unable to provide informed consent and/or those not meeting all eligibility criteria were excluded from the study. Patients eligible for the study underwent pre-test counseling conducted by eight physicians, followed by the administration of informed consent. Buccal samples were collected using OraCollect•DNA (DNA Genotek) under the supervision of trained clinic staff.

**Table 1:** List of drugs, genes, and variants included in the pharmacogenomics panel.

| Gene | Allele | Variant | Allele Functional Status | Drugs of Interest |
| --- | --- | --- | --- | --- |
| CYP2C9 | *2 | rs1799853 | Decreased | Celecoxib |
|  | *3 | rs1057910 | Decreased |  |
| CYP2C19 | *2 | rs4244285 | None | Amitriptyline<br>Citalopram<br>Clomipramine<br>Clopidogrel<br>Doxepin<br>Escitalopram<br>Imipramine<br>Lansoprazole<br>Omeprazole<br>Sertraline<br>Voriconazole |
|  | *3 | rs4986893 | None |  |
|  | *17 | rs12248560 | Increased |  |
| CYP2D6 | *2 | rs16947, rs1135840 | Normal | Amitriptyline<br>Aripiprazole<br>Atomoxetine<br>Clomipramine<br>Codeine<br>Doxepin<br>Flecainide<br>Fluvoxamine<br>Haloperidol<br>Imipramine<br>Metoprolol |
|  | *3 | rs35742686 | None |  |
|  | *4 | rs3892097, rs1065852 | None |  |
|  | *5 | NA (Deletion) | None |  |
|  | *6 | rs5030655 | None |  |
|  | *8 | rs5030865 | None | Nortriptyline<br>Ondansetron<br>Paroxetine<br>Propafenone<br>Tamoxifen<br>Tramadol<br>Venlafaxine<br>Zuclopenthixol |
|  | *9 | rs5030656 | Decreased |  |
|  | *10 | rs3892097, rs1065852 | Decreased |  |
|  | *14 | rs5030865 | None |  |
|  | *21 | rs72549352 | None |  |
|  | *29 | rs59421388 | Decreased |  |
|  | *31 | rs267608319 | None |  |
|  | *35 | rs769258 | Normal |  |
|  | *36 | NA | None |  |
|  | *41 | rs28371725 | Decreased |  |
| SLCO1B1 |  | rs4149056 | Decreased | Simvastatin |
| HLA-B | *58:01 |  | High-risk allele | Allopurinol |

#### c) Sample Processing

All collected samples were sent to the central laboratory, Raffles Diagnostica Lab, located at Raffles Hospital. Courier service was arranged twice a week for sample delivery to NalaGenetics’ Laboratory for testing.

We utilized the Nala RxReady™ panel to process the buccal samples (∼1mL) in batches, with each batch containing three patient samples. Nala RxReady™ is a multi-gene qPCR-based panel of 21 allele variants (Table 1), comprising five pharmacogenes - CYP2C9, CYP2C19, CYP2D6, SLCO1B1, and HLA-B*58:01. Genomic DNA was extracted using the Monarch® Genomic DNA Purification Kit. Subsequently, all DNA samples were analyzed using the Bio-Rad CFX96 IVD Touch™ Real-Time PCR Detection System, following the manufacturer’s instructions. The generated files were then imported into the companion software, Nala Clinical Decision Support™ (Nala CDS™), for in-depth analysis encompassing variant genotyping, diplotype determination, and phenotype translation. The clinical recommendations generated by the software were derived from comprehensive annotations available in Clinical Pharmacogenetics Implementation Consortium (CPIC), Dutch Pharmacogenomics Working Group (DPWG), Canadian Pharmacogenomics Network for Drug Safety (CPNDS), and regulatory bodies including the US Food and Drug Administration (FDA), Pharmaceuticals and Medical Devices Agency (PMDA), Swissmedic, and European Medicines Agency (EMA) (Kothary et al., 2021).

#### d) Post-test Report

Given the high level of integration of clinical workflows with the EMR system, PGx results including genotypes, phenotypes, and follow-up recommendations were seamlessly integrated into the EMR system to facilitate the study. As depicted in Figure 2 below, a summary of patients’ PGx results automatically generates when their PGx results became available for the first time. This summary can also be accessed through the “Patient Alert” tab. Drugs with follow-up recommendations were prioritized and prominently displayed on the page. The full report (Supplementary Material S1), containing clinical recommendations, can be retrieved by selecting the recommendation hyperlink to guide prescribing.

**Figure 2:**
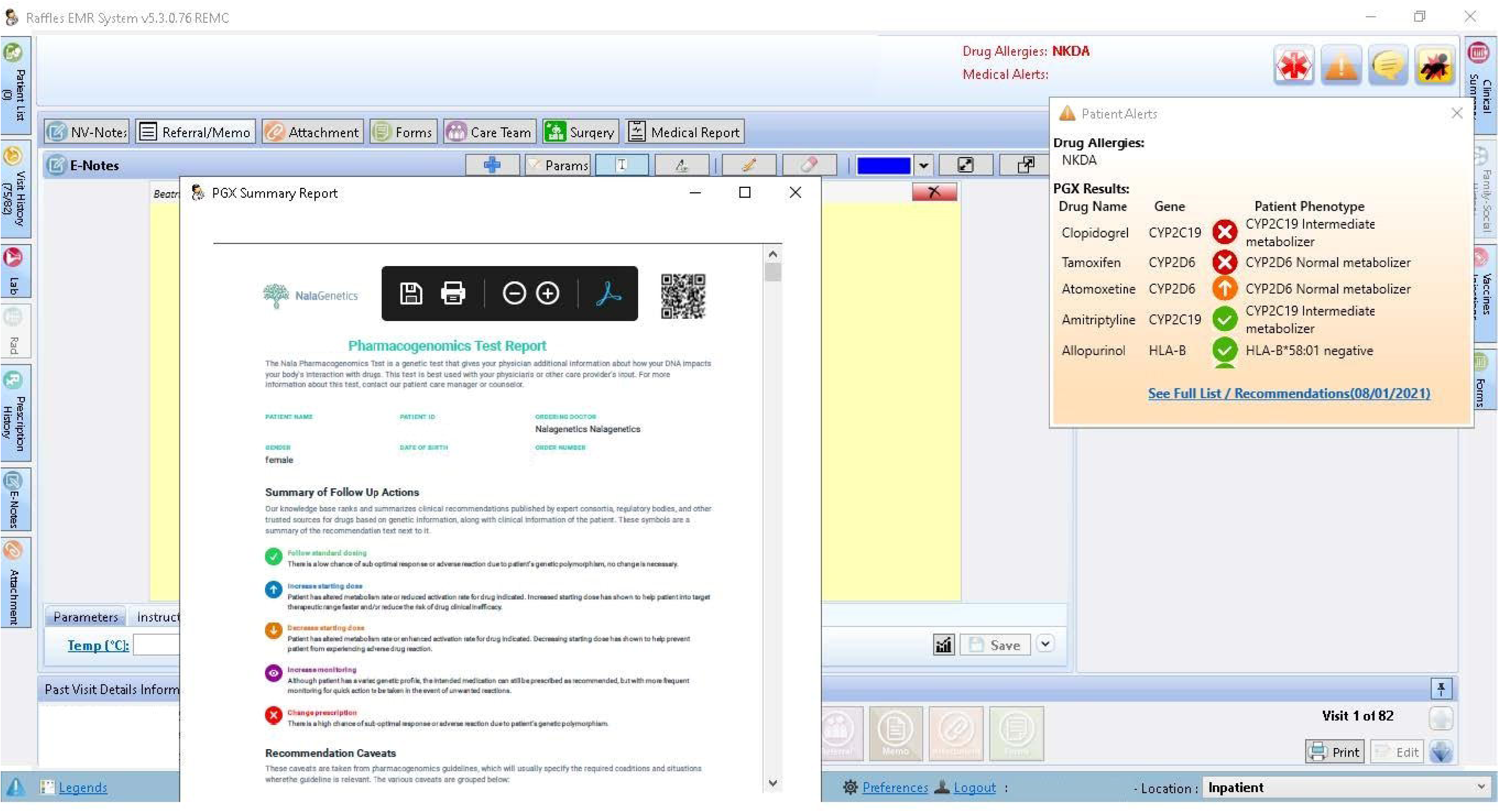
**Screenshot of the EMR interface. The summary comprises the drug names, gene list, phenotypes, follow-up recommendation symbols and a hyperlink to access the full report.**

All participating physicians were also granted access to the Nala CDS™ Platform, enabling them to view and release PGx reports for patients (Figure 3). Patients can then access their PGx results through the Nalagenetics’ Mobile application (Figure 4). Post-test counseling was offered by the physician who ordered the test during the next follow-up visits or in response to patients’ requests.

**Figure 3:**
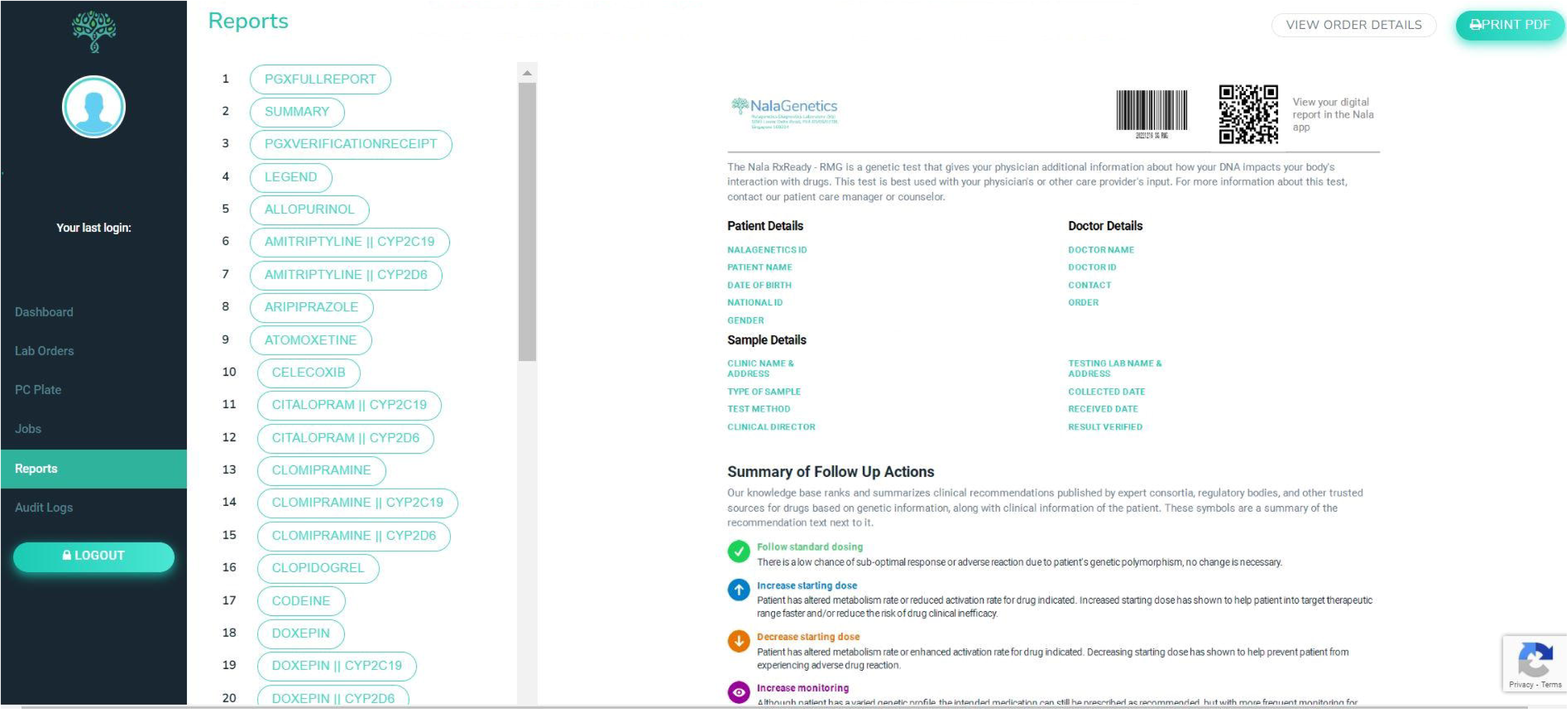
**User interface on Nala CDS Platform.**

**Figure 4:**
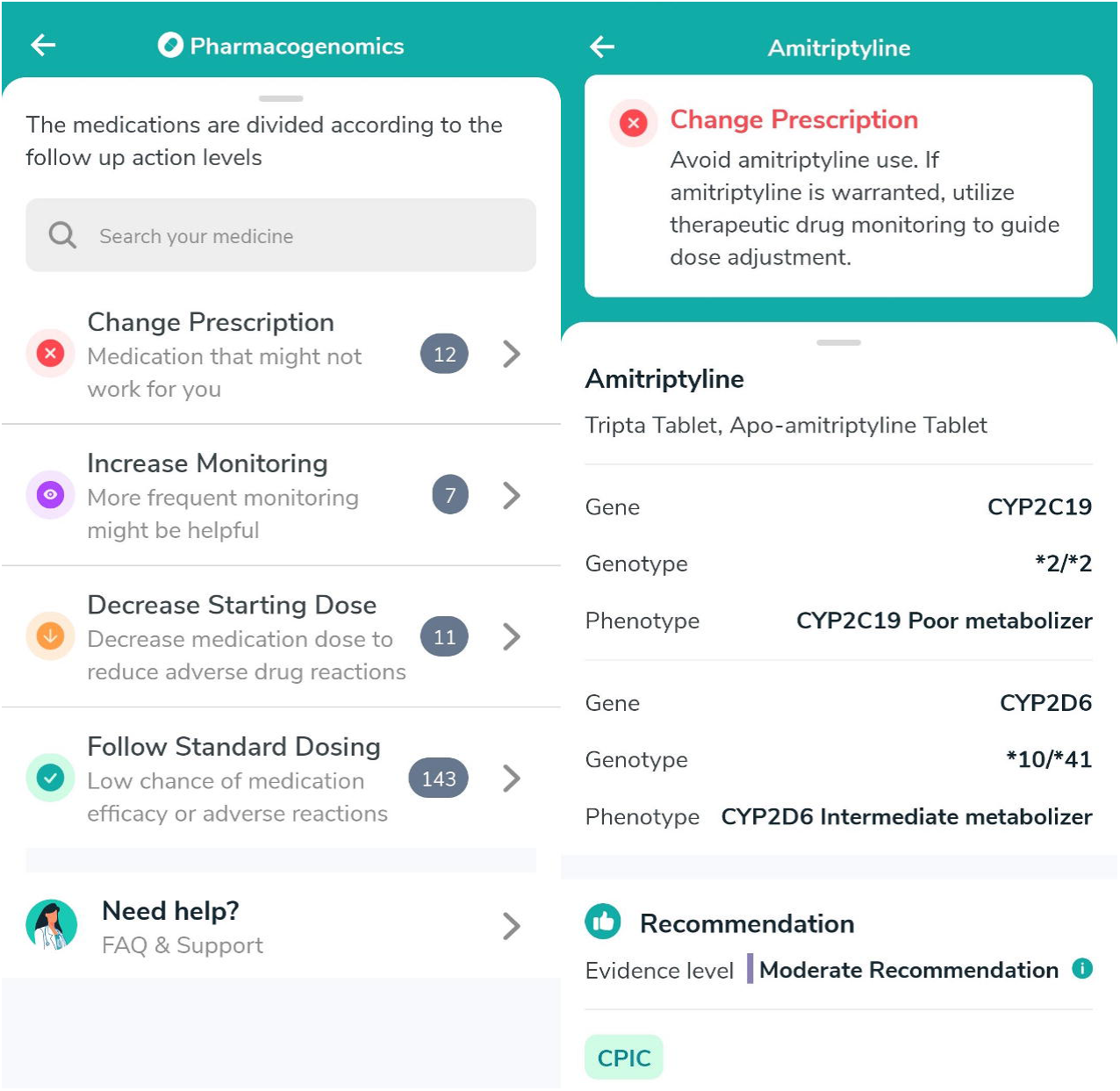
**Mobile app interface for PGx report access.**

### Data Collection

Surveys were administered to both study participants and clinicians at different intervals. All study participants completed two surveys. At enrollment, they completed a survey on their perceptions of PGx testing and medical history. At 3 months post enrollment, all study participants completed a survey providing an account on their PGx testing experience and their perceived psychological impact of receiving the PGx report. On the other hand, all clinicians completed one survey at 3 months post enrollment, which provided their experience on patient recruitment, test ordering, and perception towards PGx testing.

All surveys were designed by the study team and subsequently reviewed by the principal investigators for face validity. Responses across all surveys were structured using a combination of multiple-choice, rating scale, and Likert scale questions, with open-ended questions for additional feedback. The survey questions developed for this study are available in the supplementary material (Supplementary Material S2).

### Planned Study Outcomes

There were two key main outcomes for this study: feasibility outcomes and clinical endpoints. Feasibility outcomes encompassed the turnaround time (TAT) of PGx testing and the overall satisfaction of patients and clinicians, assessed using self-administered questionnaires. Clinical endpoints included the number of patients with actionable PGx variants.

### Sample Size Calculation and Statistical Analysis

This feasibility study aimed to enroll a sample size of 200 patients, determined based on the testing capacity provided by NalaGenetics Pte Ltd. Descriptive statistics, including mean, median, range, standard deviation, and frequency, were employed to summarize the central tendency and spread of continuous variables. Proportions were used to present statistics for categorical variables.

## Results

### Patient Recruitment and Demographics

Out of the 1428 eligible patients, 224 patients (15.7%) were recruited by 8 clinicians over an 11-months period. One patient dropped out, and one was lost to follow-up during resampling, resulting in an analyze sample of 222 patients (Figure 5). The study included 222 patients, comprising 126 females and 96 males, with an average (SD) age of 43 (10.9) years. Majority of the participants were Chinese (61.3%) by racial/ethnic origin, followed by other Asians (18.9%), Indian (9.5%), Malay (5.9%), and non-Asians (4.5%) (Table 2). Slightly over half of the patients (50.9%) self-reported to be using at least one medication.

**Figure 5:**
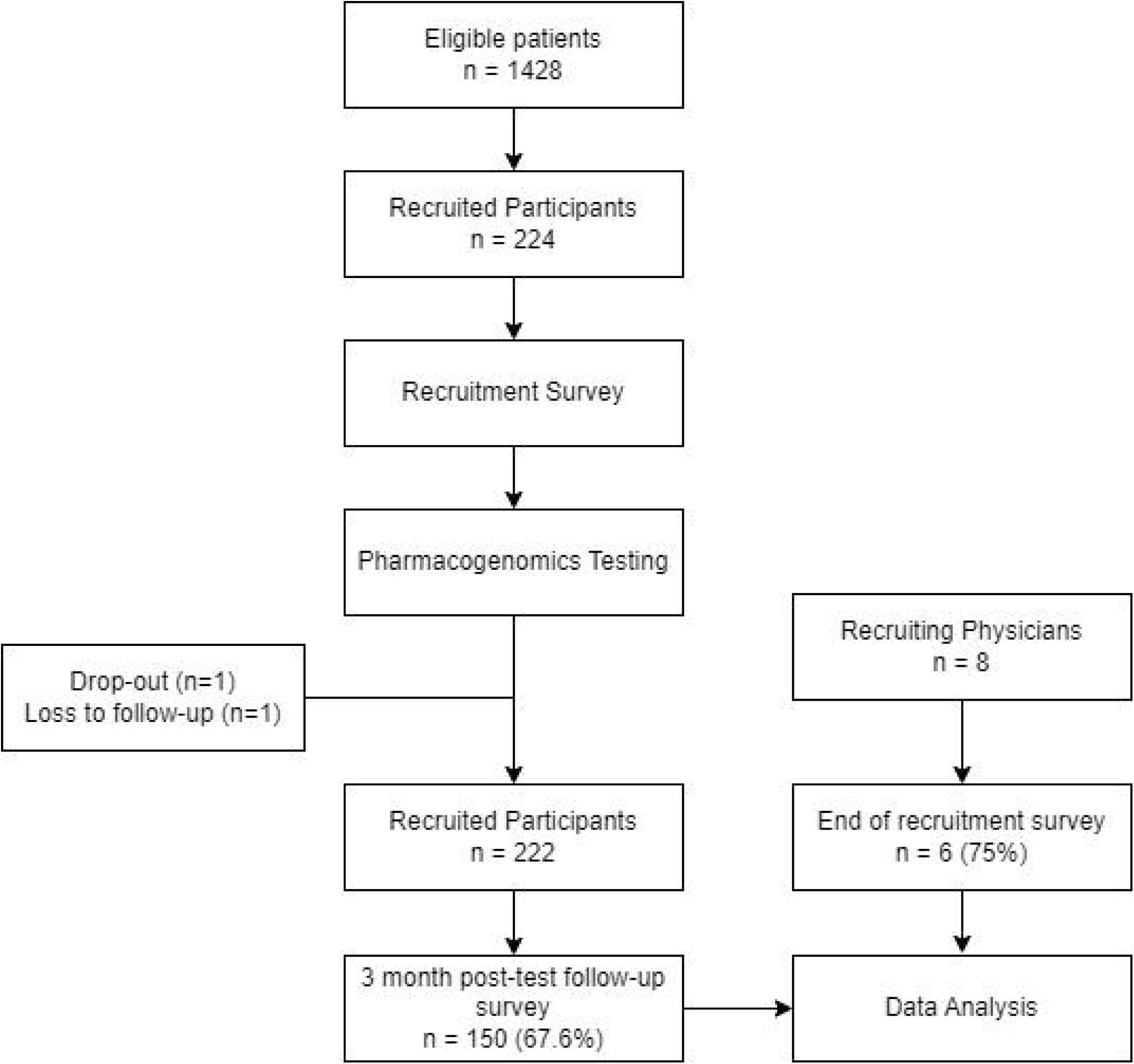
**Flowchart of study participants and recruiting physicians.**

**Table 2:**
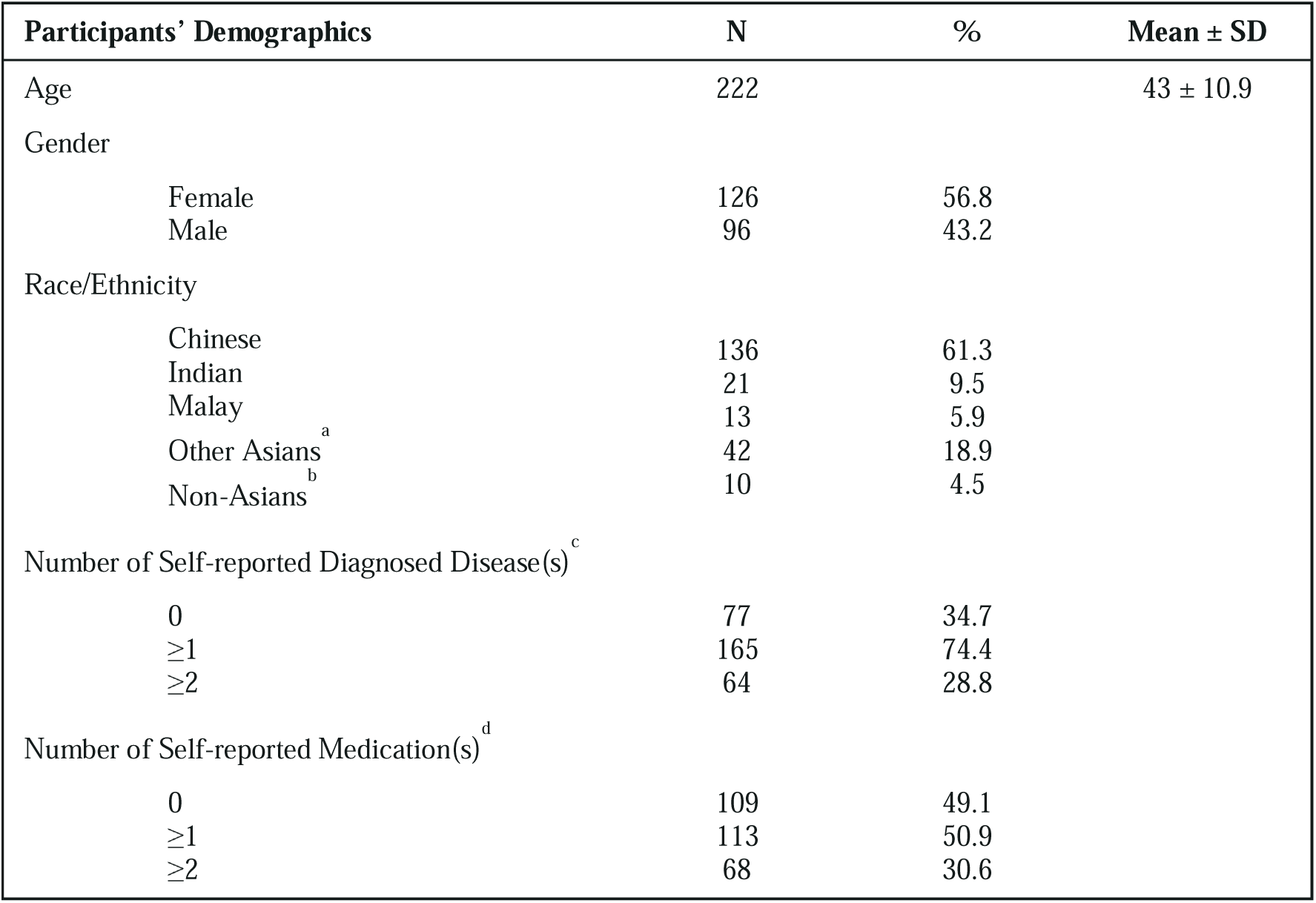
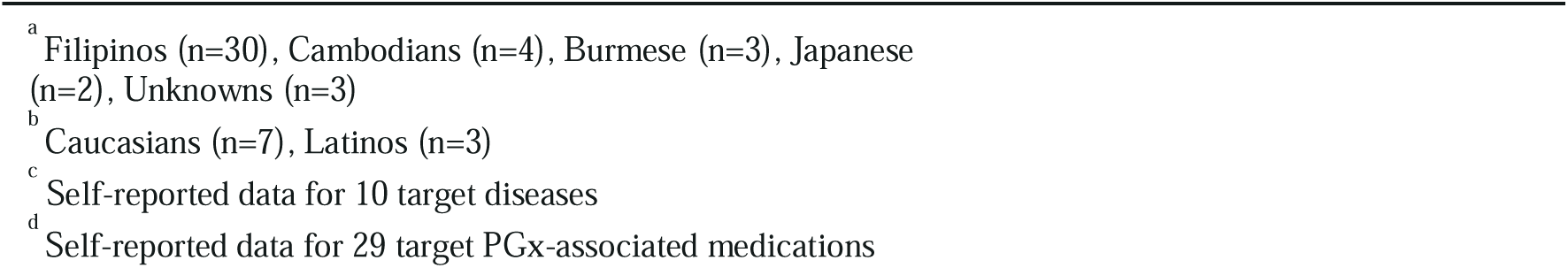
Demographics of recruited participants.

| Participants' Demographics | N | % | Mean ± SD |
| --- | --- | --- | --- |
| Age | 222 | | 43 ± 10.9 |
| Gender |  |  |  |
| Female | 126 | 56.8 |  |
| Male | 96 | 43.2 |  |
| Race/Ethnicity |  |  |  |
| Chinese | 136 | 61.3 |  |
| Indian | 21 | 9.5 |  |
| Malay | 13 | 5.9 |  |
| Other Asians<sup>a</sup> | 42 | 18.9 |  |
| Non-Asians<sup>b</sup> | 10 | 4.5 |  |
| Number of Self-reported Diagnosed Disease(s)<sup>c</sup> |  |  |  |
| 0 | 77 | 34.7 |  |
| $\geq 1$ | 165 | 74.4 | |
| $\geq 2$ | 64 | 28.8 | |
| Number of Self-reported Medication(s)<sup>d</sup> |  |  |  |
| 0 | 109 | 49.1 |  |
| $\geq 1$ | 113 | 50.9 | |
| $\geq 2$ | 68 | 30.6 | |

<sup>a</sup> Filipinos (n=30), Cambodians (n=4), Burmese (n=3), Japanese (n=2), Unknowns (n=3)
<sup>b</sup> Caucasians (n=7), Latinos (n=3)
<sup>c</sup> Self-reported data for 10 target diseases
<sup>d</sup> Self-reported data for 29 target PGx-associated medications

### Phenotype Analysis

The phenotype results of the patients were summarized in Table 3. Normal metabolizers of the CYP2D6 genes comprised 47–69% of the study population, while intermediate metabolizer phenotypes of the CYP2D6 gene were the next most prevalent among study participants, constituting a minimum of 30% in all ethnic groups except for Indians (19%). Five participants (2.3%) had uncertain results for the CYP2D6 gene, including one likely intermediate metabolizer and four with indeterminate phenotypes. For the CYP2C19 gene, ultrarapid metabolizer phenotypes were more prevalent among the non-Asian population (50%), while intermediate metabolizer phenotypes were more common among Asians, particularly among Indians (71.4%). Normal metabolizer was the most common phenotype among the study population for the CYP2C9 gene, with Indians (19.1%) and non-Asians (20%) showing a slightly higher prevalence of intermediate metabolizers. Similarly, the most common phenotype observed for the SLCO1B1 gene was normal function. However, Asians (excluding Indians) were more likely to have decreased or poor function phenotypes compared to non-Asians. Lastly, more than 80% of the study population did not carry the HLA-B*58:01 allele. This allele was more frequently observed among Chinese (18.4%), Malay (15.4%), and other Asians (11.9%), while none of the Indians and non-Asians carried it.

**Table 3:** Phenotype frequencies, presented by genes and ethnicities.

| Gene | Phenotype | Chinese<br>(n = 136) |  | Indian<br>(n = 21) |  | Malay<br>(n = 13) |  | Other Asians<br>(n = 42) |  | Non-Asians<br>(n = 10) |  |
| --- | --- | --- | --- | --- | --- | --- | --- | --- | --- | --- | --- |
|  |  | Total | Percentage (%) | Total | Percentage (%) | Total | Percentage (%) | Total | Percentage (%) | Total | Percentage (%) |
| CYP2D6 | Ultrarapid Metabolizer | 1 | 0.74 | 1 | 4.76 | 0 | 0.00 | 0 | 0.00 | 0 | 0.00 |
|  | Normal Metabolizer | 65 | 47.79 | 14 | 66.67 | 9 | 69.23 | 24 | 57.14 | 6 | 60.00 |
|  | Intermediate Metabolizer | 66 | 48.53 | 4 | 19.05 | 4 | 30.77 | 17 | 40.48 | 3 | 30.00 |
|  | Likely Intermediate Metabolizer | 1 | 0.74 | 0 | 0.00 | 0 | 0.00 | 0 | 0.00 | 0 | 0.00 |
|  | Poor Metabolizer | 1 | 0.74 | 0 | 0.00 | 0 | 0.00 | 1 | 2.38 | 1 | 10.00 |
|  | Indeterminate | 2 | 1.47 | 2 | 9.52 | 0 | 0.00 | 0 | 0.00 | 0 | 0.00 |
| CYP2C19 | Ultrarapid Metabolizer | 1 | 0.74 | 1 | 4.76 | 0 | 0.00 | 1 | 2.38 | 5 | 50.00 |
|  | Normal Metabolizer | 45 | 33.09 | 4 | 19.05 | 6 | 46.15 | 13 | 30.95 | 3 | 30.00 |
|  | Intermediate Metabolizer | 74 | 54.41 | 15 | 71.43 | 6 | 46.15 | 22 | 52.38 | 2 | 20.00 |
|  | Poor Metabolizer | 16 | 11.76 | 1 | 4.76 | 1 | 7.69 | 6 | 14.29 | 0 | 0.00 |
| CYP2C9 | Normal Metabolizer | 129 | 94.85 | 17 | 80.95 | 12 | 92.31 | 40 | 95.24 | 8 | 80.00 |
|  | Intermediate Metabolizer | 7 | 5.15 | 4 | 19.05 | 1 | 7.69 | 2 | 4.76 | 2 | 20.00 |
|  | Poor Metabolizer | 0 | 0.00 | 0 | 0.00 | 0 | 0.00 | 0 | 0.00 | 0 | 0.00 |
| SLCO1B1 | Normal Function | 104 | 76.47 | 18 | 85.71 | 9 | 69.23 | 31 | 73.81 | 9 | 90.00 |
|  | Decreased Function | 29 | 21.32 | 3 | 14.29 | 3 | 23.08 | 11 | 26.19 | 1 | 10.00 |
|  | Poor Function | 3 | 2.21 | 0 | 0.00 | 1 | 7.69 | 0 | 0.00 | 0 | 0.00 |
| HLA-B*58:01 | Positive | 25 | 18.38 | 0 | 0.00 | 2 | 15.38 | 5 | 11.90 | 0 | 0.00 |
|  | Negative | 111 | 81.62 | 21 | 100.00 | 11 | 84.62 | 37 | 88.10 | 10 | 100.00 |

Ninety-five percent of the patients had at least one clinically actionable variant, with 71.2% of the patients had at least two clinically actionable variants, and 27.5% of the patients had at least three clinically actionable variants. Based on self-reported information, 50.9% (n = 113) of our recruited patients have taken at least 1 of the 29 studied drugs, with 21.2% (n = 47) of them receiving at least one clinically actionable recommendation based on their PGx results. This included a change in prescription (26%), an increase or decrease in the starting dose (8%), and an increase in monitoring (66%).

### Turnaround Time Analysis in PGx testing

The expected TAT from sample receipt to report generation is five business days. Of the patient samples, 93% (n = 206) met the TAT requirement of five business days, with TAT ranging from 2 to 7 business days. Among the sixteen samples (7%) with TAT more than five business days, eleven were delayed due to operational issues, while five were delayed due to technical limitations.

### Experience of PGx testing at 3 months

The 3-month post-test follow-up survey explored patients’ experience with PGx testing and the psychological impact of receiving the PGx report. The response rate for the survey was 67.6% (n = 150). As summarized in Table 4, ninety-four percent (n = 141) of respondents reported satisfaction with their PGx testing experiences, with 137 indicating they would recommend PGx testing to family or friends. Upon receiving the results, the majority of the participants (70%) felt relieved and happy. Thirty-one patients (20.7%) received post-test counseling from their ordering physicians, and eight expressed a desire to seek additional information online. Additionally, more than 70% of participants reported feeling more confident in taking their prescribed medication.

**Table 4:**
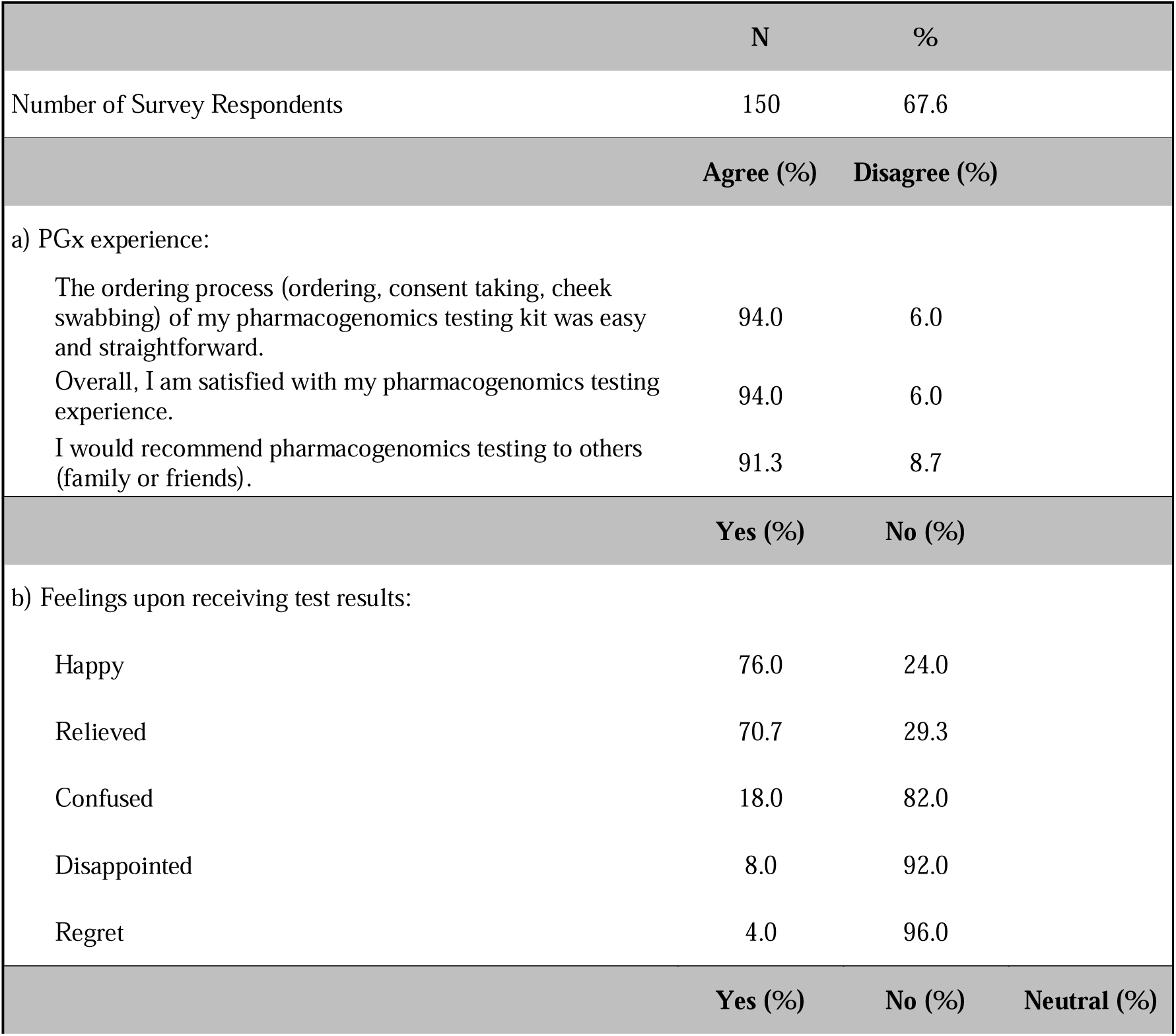
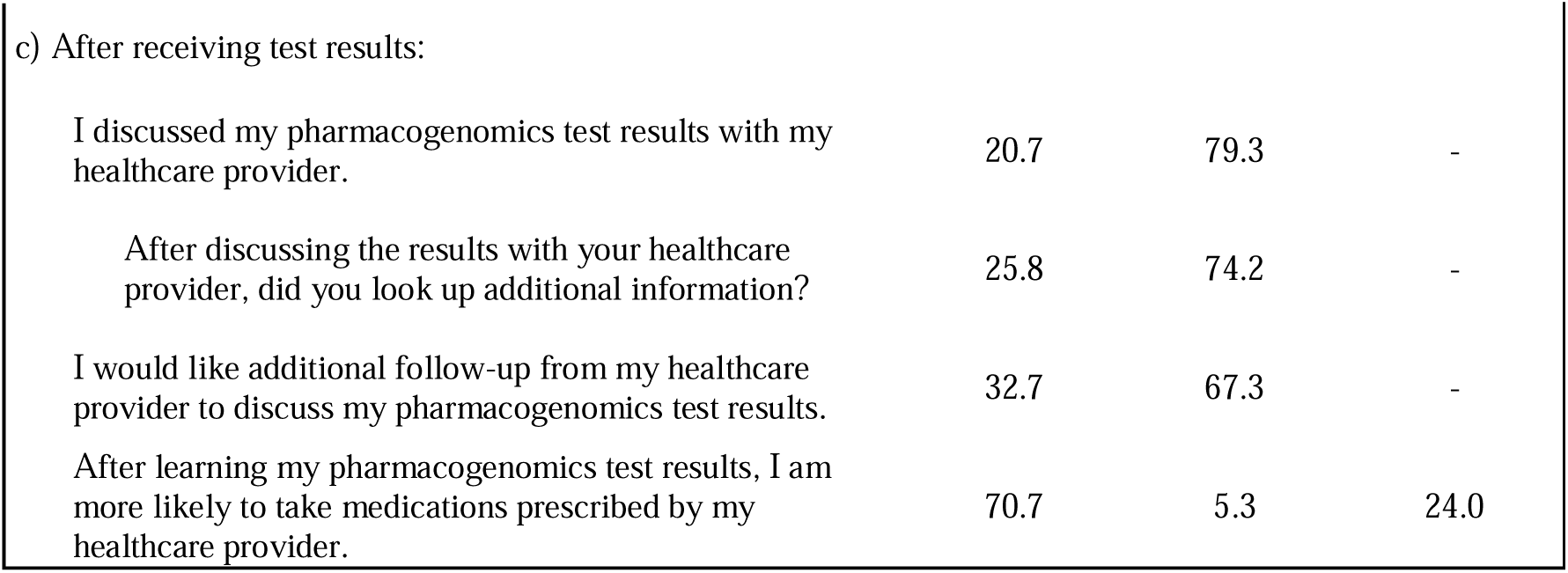
Summary of the 3-months post-test follow-up survey responses.

|  | N | % |  |
| --- | --- | --- | --- |
| Number of Survey Respondents | 150 | 67.6 |  |
|  | Agree (%) | Disagree (%) |  |
| a) PGx experience: |  |  |  |
| The ordering process (ordering, consent taking, cheek swabbing) of my pharmacogenomics testing kit was easy and straightforward. | 94.0 | 6.0 |  |
| Overall, I am satisfied with my pharmacogenomics testing experience. | 94.0 | 6.0 |  |
| I would recommend pharmacogenomics testing to others (family or friends). | 91.3 | 8.7 |  |
|  | Yes (%) | No (%) |  |
| b) Feelings upon receiving test results: |  |  |  |
| Happy | 76.0 | 24.0 |  |
| Relieved | 70.7 | 29.3 |  |
| Confused | 18.0 | 82.0 |  |
| Disappointed | 8.0 | 92.0 |  |
| Regret | 4.0 | 96.0 |  |
|  | Yes (%) | No (%) | Neutral (%) |
| c) After receiving test results: |  |  |  |
| I discussed my pharmacogenomics test results with my healthcare provider. | 20.7 | 79.3 | - |
| After discussing the results with your healthcare provider, did you look up additional information? | 25.8 | 74.2 | - |
| I would like additional follow-up from my healthcare provider to discuss my pharmacogenomics test results. | 32.7 | 67.3 | - |
| After learning my pharmacogenomics test results, I am more likely to take medications prescribed by my healthcare provider. | 70.7 | 5.3 | 24.0 |

### Perception of PGx testing: Participants

In the self-administered survey conducted at recruitment, we explored participants’ perceptions of PGx testing. The majority of participants (43%) initially learned about PGx testing from their family and friends. The top two considerations for patients deciding to participate in PGx testing were “Helpfulness in optimizing my treatment” (53.6%) and “Self-curiosity” (53.2%). In contrast, “Out-of-pocket cost” (24.8%) was identified as the least important factor for patients considering PGx testing.

### Perception of PGx testing: Clinicians

The end-of-recruitment survey for clinicians garnered valuable feedback on the recruitment process. Firstly, the extended follow-up period was identified as a deterrent to potential patient enrollment. Additionally, the challenge of reviewing test reports with patients, especially given the extensive panel covering 29 drugs, was recognized due to physicians’ busy schedules. Notably, 67% of physicians indicated a preference for using an EMR-integrated module over a separate CDS platform. Regarding their perception of PGx testing, the top two considerations for physicians when adopting PGx testing were the reasons for testing and the associated cost. Furthermore, clinicians highlighted the need for clearer legal regulations regarding PGx testing and insurance coverage for patients to enhance future adoption of PGx testing.

## Discussion

This is one of the first studies that explores the feasibility of implementing preemptive PGx testing in an Asian primary care setting. Almost all (95%) analyzed patients carried at least one clinically actionable variant using our multigene panel, suggesting the potential clinical benefits of preemptive PGx testing. This finding aligns with other studies (Smith et al., 2022; Swen et al., 2023; Van Driest et al., 2014) that were conducted in the US and in Europe. Our study also reveals that over half of the patients have taken or are currently taking at least one PGx-associated medication, with one out of 5 receiving follow-up recommendations, including changes in prescription, dosage adjustments, and increased monitoring. Our findings align with the findings of another study conducted in the Netherlands (Bank et al., 2019), where one out of 5 new prescriptions within the primary care setting were associated with actionable PGx recommendations. Collectively, these findings emphasize the potential of PGx testing to optimize patient care and underscore its relevance in personalized medicine.

We have also observed significant variations in drug metabolizing phenotypes among different ethnic groups within our study population. It is observed that over 30% of the study population, excluding Indians (17%), may exhibit the CYP2D6 Intermediate Metabolizer phenotype, thereby warranting actionable recommendations for medications such as antidepressants, tamoxifen, and flecainide. Given the high prevalence of CYP2C19 intermediate metabolizer phenotypes among Asians in Singapore, particularly within Indians, over 70% of the Indian population may necessitate an alternative antiplatelet therapy to clopidogrel. Furthermore, approximately 50% of non-Asians may exhibit the CYP2C19 Ultrarapid Metabolizer phenotype, hence requiring dosage adjustment or alternative therapies for specific antidepressants, proton pump inhibitors, and voriconazole. Asians, excluding Indians, also demonstrate a higher likelihood of developing side effects to Simvastatin (SLCO1B1) and Allopurinol (HLA-B* 58:01) compared to non-Asians (Whirl-Carrillo et al., 2021).

We have also observed a low psychological impact to patients on the return of PGx testing, as evidenced by the majority of participants feeling relieved and happy upon receiving their results. Such observation is similar to findings from other studies (Haga et al., 2014; Jameson et al., 2021) that PGx testing reports do not cause major psychological impact. It is important to acknowledge that PGx testing results can present challenges in interpretation due to their inherent complexity, particularly for patients with limited knowledge of genetics. This may prompt patients to seek further information despite having received post-testing counseling. Nonetheless, for PGx testing to effectively benefit both the patient and future healthcare providers, it is crucial for patients to fully comprehend their test results and the test’s purpose through pre- and post-test counseling. This ensures that patients can confidently share their PGx results with other healthcare providers, thereby facilitating personalized care (Haga et al., 2014).

It was interesting to observe a differing perspective between clinicians and patients regarding the cost of PGx testing, with patients having fewer concern about paying out-of-pocket costs for PGx testing. This finding contrasts with the prevailing notion that cost has consistently acted as a perceived barrier to PGx testing (Chan et al., 2017; Frigon et al., 2019). In this context, it is plausible that patients tend to prioritize the direct benefits of PGx testing, while clinicians consider factors such as clinical evidence, cost-effectiveness, and the potential financial strain on their patients. Given that the willingness to bear out-of-pocket costs for PGx testing is significantly influenced by individuals’ financial circumstances (Bielinski et al., 2017), it is noteworthy that this study was conducted in a private healthcare setting, where patients visiting private healthcare settings in Singapore might have a higher socioeconomic background.

Implementation of PGx testing in clinical practice is a complex process which encompasses PGx testing, EMR integration, and clinical impact of PGx (Giri et al., 2019). This study was also designed to explore and address some of these key considerations as part of the implementation process. To date, the sole mandatory PGx test in Singapore is genotyping for the HLA-B*15:02 allele before initiating carbamazepine (HSA, 2013). Limited studies described their experience with implementing PGx testing in healthcare settings in Singapore, with one describing the feasibility to implement preemptive PGx implementation in a primary care setting with all genotyped patients have at least one actionable PGx variants (Smith et al., 2022). In our study, the multigene panel was curated specifically to include alleles that were pertinent to Asian populations with a minor allele frequency >1% based on East Asians, South Asians, and Europeans obtained from the PharmGKB database (Kothary et al., 2021). The expected TAT of PGx testing was set at 5 business days because a long TAT may potentially lead to treatment delays. In this study, we have observed that >90% successfully met the targeted TAT. While meeting the expected TAT for PGx testing is crucial, conducting PGx testing preemptively may serve as a solution by identifying genetic factors that influence drug responses in advance. EMR integration plays a pivotal role in the successful implementation of PGx into healthcare systems (Giri et al., 2019), with majority of our clinicians (67%) preferred the usage of an EMR-integrated module over a standalone CDS platform. By seamlessly incorporating PGx results into EMR systems, this facilitates quick retrieval and display of relevant PGx data where healthcare professionals can access and utilize genetic information to make more informed and personalized decisions on patients’ medication management. Furthermore, PGx data becomes readily available for future clinical encounters, further enhancing the quality of patient care. Empowering patients in their healthcare decisions is also a crucial aspect of personalized medicine, and providing a dedicated mobile app would allow patients to gain direct access to their PGx results, thus allowing them to participate in their health management. Patients can easily review their PGx data, understand how it impacts their medication responses, and explore recommendations for personalized treatment. This encourages patients to be more proactive in discussions with their healthcare providers and to ask informed questions about their prescriptions, thus promoting shared decision-making.

There are major clinical implications with the findings of our study. We experience a low recruitment rate, with only a fraction of eligible patients enrolling in the study. Despite the potential benefits of PGx testing in optimizing medication therapy and improving patient outcomes, the challenges associated with recruitment highlight barriers that need to be addressed. For example. many patients were unfamiliar with PGx testing and its potential to guide medication selection and dosing. Therefore, efforts to increase awareness and education about the benefits of PGx testing are essential for successful implementation of PGx testing in the outpatient setting (Kabbani et al., 2023). Additionally, the observed emphasis on the need for clearer guidelines and insurance coverage to promote wider adoption of PGx testing in primary and secondary care aligns with the expected priorities in this field (Chan et al., 2017). While two major consortia, CPIC and DPWG provide valuable guidance, disparities in clinical recommendations arise from their distinct methodologies (Krebs & Milani, 2019). Furthermore, the absence of standardization in reporting formats presents a challenge (Lanting et al., 2020). Hence, a concerted effort is imperative to effectively address these issues. Furthermore, despite the growing interest in and the decreasing cost of PGx testing, cost remains a significant barrier to its implementation (Chan et al., 2017; Frigon et al., 2019). Therefore, expanding insurance coverage for PGx testing may serve as a key solution to address this barrier effectively. Previous studies have shown that patients are more inclined to undergo PGx testing when the costs are fully or partially covered by insurance (Bielinski et al., 2017; Gibson et al., 2017; Liko et al., 2020). We encourage that all stakeholders, including third-party administrators, policy makers, clinicians and PGx service providers, to jointly and regularly review existing evidence, in order to continue to establish clearer testing guidelines and expand insurance coverage for the wider adoption of PGx testing (Haidar et al., 2022; Keeling et al., 2019).

There are several strengths and weaknesses in this study. This innovative research serves as an encouraging step toward the clinical implementation of pre-emptive PGx testing in the outpatient setting in Asia, by laying the foundation for future research that can encompass a wider healthcare setting, such as tertiary hospitals. Furthermore, this study involved both primary and secondary care settings, allowing for a more comprehensive and integrated view of implementing PGx testing. One notable limitation of this study is its focus on private healthcare settings, which may restrict the generalizability of the findings to larger and more diverse populations.

## Conclusion

It is highly feasible to implement a preemptive PGx testing in outpatient clinics within private healthcare settings in Singapore. This study establishes a framework for the clinical integration of PGx testing, encompassing key aspects such as testing protocols, training, education, EMR integration, and the potential clinical impact. Our early findings emphasize the clinical benefits of PGx testing, identifying a high prevalence of clinically actionable variants in almost all patients who were being genotyped with a clinically relevant multi-gene panel. Notably, differing perspectives between patients and clinicians on the cost as a barrier to PGx testing were observed, with cost being the least important factor for patients. We encourage that all stakeholders, including third-party administrators, policy makers, clinicians and PGx service providers, to jointly and regularly review existing evidence, and establish clearer testing guidelines and expand insurance coverage for the wider adoption of PGx testing.

## Supporting information

Supplementary Material S1: Sample pharmacogenomics report

Supplementary Material S2: Survey Questions

## Data Availability

The datasets generated and/or analysed during the current study are not publicly available due to ethical and privacy reasons but are available from the corresponding author on reasonable request.

## List of abbreviations

ADE: Adverse Drug Event
CDS: Clinical Decision Support
CPIC: Clinical Pharmacogenetics Implementation Consortium
CPNDS: Canadian Pharmacogenomics Network for Drug Safety
CYP: Cytochrome P450
DPWG: Dutch Pharmacogenomics Working Group
EMA: European Medicines Agency
EMR: Electronic Medical Record
FDA: Food and Drug Administration
HLA: Human Leukocyte Antigens
PharmGKB: Pharmacogenomics KnowledgeBase
PGx: Pharmacogenomics
PMDA: Pharmaceuticals and Medical Devices Agency
qPCR: Qualitative Polymerase Chain Reaction
RMG: Raffles Medical Group
TAT: Turnaround Time

## Declarations

### Ethics approval and consent to participate

This study was approved by Raffles Hospital Institutional Review Board under reference number 2017/007.

All necessary patient/participant consent has been obtained and the appropriate institutional forms have been archived, and that any patient/participant/sample identifiers included were not known to anyone outside the research group so cannot be used to identify individuals.

### Consent for publication

Not applicable.

### Authors’ contributions

Author FN contributed to the design and implementation of the research, to the collection and analysis of the results, and to the writing of the manuscript. Author RV contributed to the implementation of the research, to the collection and analysis of the results and to the writing of the manuscript. Author AI, LS, ML, MW, AW, SK, and AC contributed to the design and implementation of the research, and to the reviewing of the manuscript.

## Acknowledgements

We would like to express our gratitude to the clinic assistants, clinic managers, nurses, pharmacists and doctors at Raffles Executive Medical Centre, Raffles Medical at Marina Bay Financial Centre, Raffles Medical at Singapore Land Tower, Raffles Diabetes and Endocrine Centre, and Raffles Neuroscience Centre for their valuable assistance in conducting this research.

## Supplementary Materials

Supplementary Material S1: Sample pharmacogenomics report

Supplementary Material S2: Survey Questions

## References

Adesta, F., Mahendra, C., Junusmin, K. I., Rajah, A. M. S., Goh, S., Sani, L., Chan, A., & Irwanto, A. (2021). Pharmacogenomics Implementation Training Improves Self-Efficacy and Competency to Drive Adoption in Clinical Practice. Frontiers in Pharmacology, 12. 10.3389/fphar.2021.684907

Bank, P. C. D., Swen, J. J., & Guchelaar, H. J. (2019). Estimated nationwide impact of implementing a preemptive pharmacogenetic panel approach to guide drug prescribing in primary care in the Netherlands. BMC Medicine, 17(1). 10.1186/s12916-019-1342-5

Bielinski, S. J., St Sauver, J. L., Olson, J. E., Wieland, M. L., Vitek, C. R., Bell, E. J., Mc Gree, M. E., Jacobson, D. J., McCormick, J. B., Takahashi, P. Y., Black, J. L., Caraballo, P. J., Sharp, R. R., Beebe, T. J., Weinshilboum, R. M., Wang, L., & Roger, V. L. (2017). Are patients willing to incur out-of-pocket costs for pharmacogenomic testing? Pharmacogenomics Journal, 17(1). 10.1038/tpj.2016.72

Chan, C. Y. W., Chua, B. Y., Subramaniam, M., Suen, E. L. K., & Lee, J. (2017). Clinicians’ perceptions of pharmacogenomics use in psychiatry. Pharmacogenomics, 18(6). 10.2217/pgs-2016-0164

Dunnenberger, H. M., Crews, K. R., Hoffman, J. M., Caudle, K. E., Broeckel, U., Howard, S. C., Hunkler, R. J., Klein, T. E., Evans, W. E., & Relling, M. V. (2015). Preemptive clinical pharmacogenetics implementation: Current programs in five us medical centers. In Annual Review of Pharmacology and Toxicology (Vol. 55). 10.1146/annurev-pharmtox-010814-124835

Frigon, M. P., Blackburn, M. È., Dubois-Bouchard, C., Gagnon, A. L., Tardif, S., & Tremblay, K. (2019). Pharmacogenetic testing in primary care practice: Opinions of physicians, pharmacists and patients. Pharmacogenomics, 20(8). 10.2217/pgs-2019-0004

Gibson, M. L., Hohmeier, K. C., & Smith, C. T. (2017). Pharmacogenomics testing in a community pharmacy: Patient perceptions and willingness-to-pay. Pharmacogenomics, 18(3). 10.2217/pgs-2016-0161

Giri, J., Moyer, A. M., Bielinski, S. J., & Caraballo, P. J. (2019). Concepts driving pharmacogenomics implementation into everyday healthcare. In Pharmacogenomics and Personalized Medicine (Vol. 12). 10.2147/PGPM.S193185

Gottesman, O., Scott, S. A., Ellis, S. B., Overby, C. L., Ludtke, A., Hulot, J. S., Hall, J., Chatani, K., Myers, K., Kannry, J. L., & Bottinger, E. P. (2013). The CLIPMERGE PGx program: Clinical implementation of personalized medicine through electronic health records and genomics-pharmacogenomics. Clinical Pharmacology and Therapeutics, 94(2). 10.1038/clpt.2013.72

Haga, S. B., Mills, R., & Bosworth, H. (2014). Striking a balance in communicating pharmacogenetic test results: Promoting comprehension and minimizing adverse psychological and behavioral response. In Patient Education and Counseling (Vol. 97, Issue 1). 10.1016/j.pec.2014.06.007

Haidar, C. E., Crews, K. R., Hoffman, J. M., Relling, M. V., & Caudle, K. E. (2022). Advancing Pharmacogenomics from Single-Gene to Preemptive Testing. In Annual Review of Genomics and Human Genetics (Vol. 23). 10.1146/annurev-genom-111621-102737

HSA. (2013, August 29). Recommendations for HLA-B*1502 genotype testing prior to initiation of carbamazepine in new patients. https://www.hsa.gov.sg/announcements/safety-alert/recommendations-for-hla-b-1502-genotype-testing-prior-to-initiation-of-carbamazepine-in-new-patients

Jameson, A., Fylan, B., Bristow, G. C., Sagoo, G. S., Dalton, C., Cardno, A., Sohal, J., & McLean, S. L. (2021). What Are the Barriers and Enablers to the Implementation of Pharmacogenetic Testing in Mental Health Care Settings? In Frontiers in Genetics (Vol. 12). 10.3389/fgene.2021.740216

Ji, Y., Skierka, J. M., Blommel, J. H., Moore, B. E., Vancuyk, D. L., Bruflat, J. K., Peterson, L. M., Veldhuizen, T. L., Fadra, N., Peterson, S. E., Lagerstedt, S. A., Train, L. J., Baudhuin, L. M., Klee, E. W., Ferber, M. J., Bielinski, S. J., Caraballo, P. J., Weinshilboum, R. M., & Black, J. L. (2016). Preemptive Pharmacogenomic Testing for Precision Medicine: A Comprehensive Analysis of Five Actionable Pharmacogenomic Genes Using Next-Generation DNA Sequencing and a Customized CYP2D6 Genotyping Cascade. Journal of Molecular Diagnostics, 18(3). 10.1016/j.jmoldx.2016.01.003

Kabbani, D., Akika, R., Wahid, A., Daly, A. K., Cascorbi, I., & Zgheib, N. K. (2023). Pharmacogenomics in practice: a review and implementation guide. In Frontiers in Pharmacology (Vol. 14). 10.3389/fphar.2023.1189976

Keeling, N. J., Rosenthal, M. M., West-Strum, D., Patel, A. S., Haidar, C. E., & Hoffman, J. M. (2019). Preemptive pharmacogenetic testing: exploring the knowledge and perspectives of US payers. Genetics in Medicine, 21(5). 10.1038/gim.2017.181

Kimpton, J. E., Carey, I. M., Threapleton, C. J. D., Robinson, A., Harris, T., Cook, D. G., DeWilde, S., & Baker, E. H. (2019). Longitudinal exposure of English primary care patients to pharmacogenomic drugs: An analysis to inform design of pre-emptive pharmacogenomic testing. British Journal of Clinical Pharmacology, 85(12). 10.1111/bcp.14100

Kothary, A. S., Mahendra, C., Tan, M., Min Tan, E. J., Hong Yi, J. P., Gabriella, G., Hui Jocelyn, T. X., Haruman, J. S., Tan, Z., Lee, C. K., Lezhava, A., Yan, B., & Irwanto, A. (2021). Validation of a multi-gene qPCR-based pharmacogenomics panel across major ethnic groups in Singapore and Indonesia. Pharmacogenomics, 22(16). 10.2217/pgs-2021-0071

Krebs, K., & Milani, L. (2019). Translating pharmacogenomics into clinical decisions: do not let the perfect be the enemy of the good. In Human genomics (Vol. 13, Issue 1). 10.1186/s40246-019-0229-z

Lanting, P., Drenth, E., Boven, L., van Hoek, A., Hijlkema, A., Poot, E., van der Vries, G., Schoevers, R., Horwitz, E., Gans, R., Kosterink, J., Plantinga, M., van Langen, I., Ranchor, A., Wijmenga, C., Franke, L., Wilffert, B., & Sijmons, R. (2020). Practical barriers and facilitators experienced by patients, pharmacists and physicians to the implementation of pharmacogenomic screening in Dutch outpatient hospital care—an explorative pilot study. Journal of Personalized Medicine, 10(4). 10.3390/jpm10040293

Liko, I., Lai, E., Griffin, R. J., Aquilante, C. L., & Lee, Y. M. (2020). Patients’ Perspectives on Psychiatric Pharmacogenetic Testing. Pharmacopsychiatry, 53(6). 10.1055/a-1183-5029

Raffles Medical Group. (2023). Annual Report 2022, Annual and Sustainability Reports. . https://www.rafflesmedicalgroup.com/wp-content/uploads/2023/04/RMG-Annual-Report-2022.pdf

Rollinson, V., Turner, R., & Pirmohamed, M. (2020). Pharmacogenomics for primary care: An overview. In Genes (Vol. 11, Issue 11). 10.3390/genes11111337

Schildcrout, J. S., Denny, J. C., Bowton, E., Gregg, W., Pulley, J. M., Basford, M. A., Cowan, J. D., Xu, H., Ramirez, A. H., Crawford, D. C., Ritchie, M. D., Peterson, J. F., Masys, D. R., Wilke, R. A., & Roden, D. M. (2012). Optimizing drug outcomes through pharmacogenetics: A case for preemptive genotyping. Clinical Pharmacology and Therapeutics, 92(2). 10.1038/clpt.2012.66

Smith, H., Dawes, M., Katzov-Eckert, H., Burrell, S., Xin Hui, S., & Winther, M. D. (2022). Improving prescribing: a feasibility study of pharmacogenetic testing with clinical decision support in primary healthcare in Singapore. Family Practice. 10.1093/fampra/cmac124

Swen, J. J., van der Wouden, C. H., Manson, L. E., Abdullah-Koolmees, H., Blagec, K., Blagus, T., Böhringer, S., Cambon-Thomsen, A., Cecchin, E., Cheung, K. C., Deneer, V. H., Dupui, M., Ingelman-Sundberg, M., Jonsson, S., Joefield-Roka, C., Just, K. S., Karlsson, M. O., Konta, L., Koopmann, R., … Rajasingam, A. (2023). A 12-gene pharmacogenetic panel to prevent adverse drug reactions: an open-label, multicentre, controlled, cluster-randomised crossover implementation study. The Lancet, 401(10374). 10.1016/S0140-6736(22)01841-4

Tsermpini, E. E., Skokou, M., Ferentinos, P., Georgila, E., Gourzis, P., Assimakopoulos, K., & Patrinos, G. P. (2020). Clinical implementation of preemptive pharmacogenomics in psychiatry: Τhe “PREPARE” study. Psychiatrike = Psychiatriki, 31(4). 10.22365/jpsych.2020.314.341

Van Driest, S. L., Shi, Y., Bowton, E., Schildcrout, J., Peterson, J., Pulley, J., Denny, J., & Roden, D. (2014). Clinically actionable genotypes among 10,000 patients with preemptive pharmacogenomic testing. Clinical Pharmacology and Therapeutics, 95(4). 10.1038/clpt.2013.229

Wang, L., Scherer, S. E., Bielinski, S. J., Muzny, D. M., Jones, L. A., Black, J. L., Moyer, A. M., Giri, J., Sharp, R. R., Matey, E. T., Wright, J. A., Oyen, L. J., Nicholson, W. T., Wiepert, M., Sullard, T., Curry, T. B., Rohrer Vitek, C. R., McAllister, T. M., St. Sauver, J. L., … Weinshilboum, R. M. (2022). Implementation of preemptive DNA sequence–based pharmacogenomics testing across a large academic medical center: The Mayo-Baylor RIGHT 10K Study. Genetics in Medicine, 24(5). 10.1016/j.gim.2022.01.022

Whirl-Carrillo, M., Huddart, R., Gong, L., Sangkuhl, K., Thorn, C. F., Whaley, R., & Klein, T. E. (2021). An Evidence-Based Framework for Evaluating Pharmacogenomics Knowledge for Personalized Medicine. Clinical Pharmacology and Therapeutics, 110(3), 563–572. 10.1002/CPT.2350

