## Supplementary figures and images for "Implementation of Pre-emptive Pharmacogenomics Testing in Outpatient Clinics in Asia (IMPT study)"

### Supplementary Material S1: Sample pharmacogenomics report

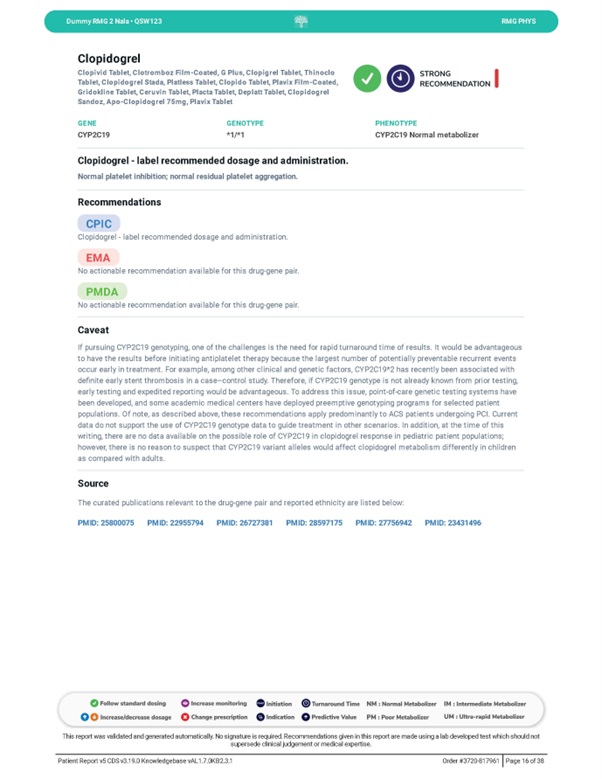
