## Supplementary Material S2: Survey Questions for "Implementation of Pre-emptive Pharmacogenomics Testing in Outpatient Clinics in Asia (IMPT study)"

**Survey 1:** Patient Recruitment Survey

| | |
|---|---|
| 1. Full Name | • [Open-ended response] |
| 2. Contact Number | • [Open-ended response] |
| 3. Email Address | • [Open-ended response] |
| 4. How old are you? (in years) | • [Open-ended response] |
| 5. In your opinion, how healthy are you right now? | • Healthy<br>• Somewhat healthy<br>• Unhealthy<br>• Very Healthy |
| 6. How interested are you in learning about genetic risk for the following conditions? (Optional)<br>a. Osteoarthritis<br>b. Rheumatoid arthritis<br>c. Asthma<br>d. Celiac disease<br>e. Ulcerative colitis<br>f. Breast cancer<br>g. Colorectal cancer<br>h. Leukemia<br>i. Lung cancer<br>j. Prostate cancer<br>k. Skin cancer<br>l. Heart disease<br>m. Blood coagulation/embolic disease<br>n. Chronic Kidney Disease<br>o. High cholesterol<br>p. Diabetes<br>q. Glaucoma<br>r. Bipolar disorder<br>s. Alzheimer's disease<br>t. Lou Gehrig's disease<br>u. Multiple sclerosis<br>v. Parkinson's disease<br>w. Obesity | • Not interested at all<br>• Somewhat uninterested<br>• Interested<br>• Very interested |
| 7. How did you first hear about genetic testing for drug response? | • Your doctor<br>• Friends and family<br>• Social media<br>• News<br>• Other companies |

| | |
|---|---|
| | • I have never heard of genetic testing for drug response<br>• Other (please specify): [Open-ended response] |
| 8. What is the reason you are seeing the doctor today? | • Routine check-up<br>• To consult on a health problem<br>• Follow-up for a chronic illness or post-operation<br>• Accompanying a patient<br>• Other (please specify): [Open-ended response] |
| 9. To what extent did the following factors play a part in your decision to be tested?<br>a. Helpfulness in optimizing my treatment<br>b. Recommendation from my healthcare provider<br>c. Recommendation from my family or friends<br>d. Self-curiosity<br>e. Out-of-pocket cost<br>f. Personal experience or stories of side effects caused by medication<br>g. Safety of personal medical and genetic data | • Not at all<br>• Very little<br>• Somewhat<br>• To a great extent |
| 10. Are you currently taking any chronic medication? | • Yes<br>• No |
| 11. Do you expect to take any daily/chronic medication in the future? | • Yes<br>• No |
| 12. Have you taken any of the following medication before? | • Allopurinol<br>• Amitriptyline<br>• Aripiprazole<br>• Atomoxetine<br>• Celecoxib<br>• Citalopram<br>• Clomipramine<br>• Clopidogrel<br>• Codeine<br>• Doxepin<br>• Escitalopram<br>• Flecainide<br>• Fluvoxamine<br>• Haloperidol |

| | |
|---|---|
| | • Imipramine<br>• Lansoprazole<br>• Metoprolol<br>• Nortriptyline<br>• Omeprazole<br>• Ondansetron<br>• Paroxetine<br>• Propafenone<br>• Sertraline<br>• Simvastatin<br>• Tamoxifen<br>• Tramadol<br>• Venlafaxine<br>• Voriconazole<br>• Zuclopenthixol |
| 13. Have you ever been diagnosed or is currently diagnosed with the following conditions? | • Type 2 Diabetes Mellitus<br>• Hypertension<br>• Gout<br>• Anxiety<br>• Stroke<br>• Rheumatoid arthritis<br>• Osteoarthritis<br>• Hyperlipidemia (high cholesterol)<br>• Ischaemic heart disease (heart attack)<br>• Major depressive disorder (depression) |
| 14. How many times have you been hospitalized (with overnight stay) within the last year? | • [Open-ended response] |
| 15. Please list the reason(s) for your hospitalization below: (Check all that apply) | • Accident, poisoning and violence<br>• Cancer<br>• Acute respiratory infection<br>• Skin infection<br>• Gastroenterological infection<br>• Heart disease<br>• Cerebrovascular diseases (including stroke)<br>• Pregnancy complications affecting foetus and newborn |

| | |
|---|---|
| | • Other (please specify): [Open-ended response] |
| 16. How many times have you visited a hospital within the last year (excluding health screening and hospitalization)? | • [Open-ended response] |
| 17. Please list the reason(s) for your hospital visit below: (Check all that apply) | • Cancer<br>• Ear, Nose & Throat Conditions (e.g., General ENT, Allergy Testing)<br>• Eye Conditions (e.g., Cataract, Refractive Surgery)<br>• Heart Conditions<br>• Neurology Conditions (e.g., Epilepsy)<br>• Pain Management (e.g., Arthritis, Spinal Pain)<br>• Skin Conditions (e.g., Skin Rash)<br>• Urology (e.g., Kidney Stones, Urinary Tract Problems)<br>• Other (please specify): [Open-ended response] |
| 18. Have your doctor told you that you experience adverse drug reaction before in your lifetime? | • Yes<br>• No<br>• I'm not sure<br>• Other (please specify): [Open-ended response] |
| 19. Have you ever felt that your medication did not work as expected? | • Yes<br>• No<br>• I'm not sure |
| 20. If yes, please list the name of medication that might have caused this: | • [Open-ended response] |
| 21. Have you suspected your direct family member or others of experiencing adverse drug reaction before in their lifetime?<br>a. Father<br>b. Mother<br>c. Sibling<br>d. Paternal Grandfather<br>e. Paternal Grandmother<br>f. Maternal Grandfather<br>g. Maternal Grandmother<br>h. Other family members | • Yes<br>• No<br>• Maybe |

| | |
|---|---|
| i. Not a family member | |
| 22. If you have answered "Yes" to the previous question, please list down what are the medications that caused the adverse drug reaction. | • [Open-ended response] |

**Survey 2:** Patient 3 Months Post-test Follow-up Survey

| | |
|---|---|
| **Section 1: My Testing Experience**<br>1. Please help us to understand your testing experience by selecting which of the following BEST describes it<br>a. The ordering process (ordering, consent taking, cheek swabbing) of my pharmacogenomics testing kit was easy and straightforward.<br>b. I find pharmacogenomics testing to be helpful to me in my healthcare decision-making at this time.<br>c. I believe pharmacogenomics testing will be helpful to me in my healthcare decision-making in the future.<br>d. I am satisfied with my decision to take the pharmacogenomics test.<br>e. Overall, I am satisfied with my pharmacogenomics testing experience. | • Strongly Disagree<br>• Somewhat Disagree<br>• Somewhat Agree<br>• Strongly Agree |
| 2. How did you learn about your pharmacogenomic test results? (Check all that apply) | • Nala Personal Health Manager Mobile app<br>• In an Email or Message<br>• In-person at a follow-up appointment scheduled with my healthcare provider specifically to view results<br>• In-person at an appointment scheduled for another reason<br>• Can't recall<br>• Other (please specify): [Open-ended response] |
| 3. What would be your preferred method of receiving your pharmacogenomic test results? | • Nala Personal Health Manager Mobile app<br>• In an Email or Message<br>• In-person at a follow-up appointment scheduled with my healthcare provider specifically to view results<br>• In-person at an appointment scheduled for another reason |

| | |
|---|---|
| | • Other (please specify): [Open-ended response] |
| 4. I have a clear understanding of my test results from pharmacogenomics testing. | • Strongly disagree<br>• Disagree<br>• Neither agree nor disagree<br>• Agree<br>• Strongly agree |
| 5. I discussed my pharmacogenomics test results with my healthcare provider. | • Yes<br>• No (if No, please skip the following 2 questions) |
| 6. My healthcare provider seemed confident in explaining the meaning of the pharmacogenomics test results to me. | • Strongly disagree<br>• Disagree<br>• Neither agree nor disagree<br>• Somewhat agree<br>• Strongly agree |
| 7. After discussing the results with your healthcare provider, did you look up additional information? | • Yes<br>• No |
| 8. I would like additional follow-up from my healthcare provider to discuss my pharmacogenomics test results. | • Yes<br>• No |
| 9. After learning my pharmacogenomics test results, I am more likely to take medications prescribed by my healthcare provider. | • Strongly disagree<br>• Disagree<br>• Neither agree nor disagree<br>• Somewhat agree<br>• Strongly agree |
| 10. After learning my pharmacogenomics test results, I feel more confident that medication(s) prescribed to me will not cause side effects and/or will help my condition, compared to past prescriptions I've received without testing. | • Strongly disagree<br>• Disagree<br>• Neutral<br>• Agree<br>• Strongly agree<br>• Not Applicable (I currently do not take medications) |
| 11. After learning my pharmacogenomic test results, I feel more validated about my medication experiences. | • Strongly disagree<br>• Disagree<br>• Neutral<br>• Agree<br>• Strongly agree |

| | |
|---|---|
| | • Not Applicable (I currently do not take medications) |
| 12. I believe that the most valuable outcome for me from pharmacogenomics testing is: | • Decreased side effects<br>• Increased medication effectiveness<br>• Decreased cost of medications<br>• Decreased cost of related healthcare services<br>• Decreased trial and error in prescribing my medications<br>• More willing to try new medication<br>• More confidence in my prescribing healthcare provider |
| **Section 2: Feelings upon receiving Test Results**<br>13. Since receiving my test results, I have felt ____ about them:<br>a. Disappointed<br>b. Confused<br>c. Regret<br>d. Relieved<br>e. Happy | • Never<br>• Rarely<br>• Occasionally<br>• Often<br>• All the time |
| 14. I discussed my pharmacogenomics test results with a family member. | • Yes<br>• No (if no, please skip the following question)<br>• I plan to do so |
| 15. After learning about my pharmacogenomics testing experience, one or more of my family members decided to pursue pharmacogenomics testing. | • Yes<br>• No<br>• Unsure |

**Survey 3:** Principal Investigator End of Recruitment Survey

| | |
|---|---|
| **Section 1: Patient on-boarding and enrolment process**<br>1. How difficult was it to convince the participants to take our PGx test? | • 1 (Easy)<br>• 2<br>• 3<br>• 4 (Difficult) |
| 2. On average, how long did it take to convince the participant to participate in the PGx testing? | • [Open-ended response] |
| 3. What were the concerns the participants had when you offered them our PGx test?<br>a. Patient does not understand what PGx testing is for even after explaining<br>b. Patient does not see the benefit of PGx testing<br>c. Patient is afraid that the PGx testis expensive OR it will cause his/her medical services to be more expensive in the future<br>d. Patient is afraid that the results will affect their lifestyle in the future<br>e. Patient is afraid that the results will affect their employability in the future<br>f. Patient is afraid that the results will affect their medical insurability in the future | • Strongly Disagree<br>• Disagree<br>• Agree<br>• Strongly Agree |
| 4. How often do the participants ask these following questions?<br>a. What is PGx test<br>b. Cost of the PGx test<br>c. When and how will they receive the test<br>d. The reason they need the test<br>e. How will the test impact them and their family members | • Never asked<br>• Seldom asked<br>• Sometimes asked<br>• Often asked<br>• Always asked |
| 5. Please list other questions the patients asked about. | • [Open-ended response] |
| 6. What were the participants' enrolled primary (initial) intention of seeing you? | Please rank the responses below from 1 (most common) to 5 (least common)<br>• To consult you about their acute conditions (e.g. fever, gastritis, headache, pain etc)<br>• and/or to obtain Medical Certificate |

| | |
|---|---|
| | • To consult you about their chronic conditions<br>• To consult you on their routine scheduled health check-up<br>• To accompany their family/friends on their medical consultation<br>• To follow-up on their recent hospitalization or medical procedure |
| 7. What are the other problems or trends observed during recruitment process? (Optional) | • [Open-ended response] |
| 8. Please let us know how we can improve the workflow in the future. (Optional) | • [Open-ended response] |
| 9. Please rank these values in terms of importance for the participant: | Please rank the responses below from 1 (most important) to 7 (least important)<br>• The reason for the test<br>• The cost of the test<br>• The implication of the test towards their lifestyle<br>• The implication of the test towards their employability<br>• The implication of the test towards their medical insurability<br>• How trustworthy the genetic testing company is<br>• How secure their genetic data will be stored |
| **Section 2: Review of the web interface (Nala Clinical Decision Support)**<br>10. The platform I used for receiving results is | • EHR-integrated module<br>• Web application provided by Nalagenetics (Nala Clinical Decision Support)<br>• Both |
| 11. Please indicate how frequently you use each of the pages listed below on your Nala CDS web application. | • Never<br>• Rarely<br>• Sometimes |

| | |
|---|---|
| a. Dashboard<br>b. Orders<br>c. Reports<br>d. Patients<br>e. Bulk Order<br>f. Profile Settings | • Often<br>• Always |
| 12. On a scale of 1- 5, how would you rate your experience of using the Nala CDS Platform? | • 1 (Poor)<br>• 2<br>• 3<br>• 4<br>• 5 (Excellent) |
| 13. On a scale of 1-10, how likely are you to recommend Nala CDS Platform (either as EHR-integrated module, standalone web app, or both) to a friend or colleague in a similar profession? | • 1 (Very unlikely)<br>• 2<br>• 3<br>• 4<br>• 5<br>• 6<br>• 7<br>• 8<br>• 9<br>• 10 (Very likely) |
| 14. What improvements would you suggest for Nala CDS Platform? | • [Open-ended response] |
| 15. In the future, would you be interested to participate in our user research on Nala CDS? | • Yes<br>• No |
| **Section 3: Feedback on Nalagenetics Operational Staff**<br>16. The operational staff is:<br>a. Helpful -takes the initiative to check if there is anything wrong<br>b. Resourceful -knows how to solve complicated and unexpected problems<br>c. Responsive -replies in a timely manner<br>d. Articulate -able to communicate effectively<br>e. Friendly -maintains good rapport with you and clinic staff | • Strongly Disagree<br>• Disagree<br>• Agree<br>• Strongly Agree |
| 17. What can be improved from the operational staff? Please fill in if you have answered "Disagree or Strongly Disagree" in any of the above questions. | • [Open-ended response] |
| 18. Please rate the following | • Strongly Disagree<br>• Disagree |

| | |
|---|---|
| a. The operational staff helped in convincing the participants to take the test<br>b. The operational staff helped ensure the workflow is smooth | • Agree<br>• Strongly Agree |
| **Section 4: Your perception on PGx testing and future PGx**<br>19. How important will these factors be for you when making a decision for patients to take a PGx test? | Please rank these factors from 1 (most important) to 8 (least important)<br>• Patient's family history<br>• Patient's current medication<br>• Familiarity with the testing company<br>• Scientific basis of the PGx test<br>• Patient's comorbidities<br>• Patient's demographics (gender, age, race)<br>• Patient's financial capability<br>• Patient's history of therapy failure or adverse drug reaction |
| 20. I plan to use PGx test in the future to:<br>a. Guide future drug selection when appropriate<br>b. Guide future dosing when appropriate<br>c. Guide future drug monitoring when appropriate<br>d. Call for pharmacy consultation<br>e. I do not plan to use PGx test anymore | • Strongly Disagree<br>• Disagree<br>• Agree<br>• Strongly Agree |
| 21. I expect to order or recommend PGx test in the next 6 months | • 1 (Very unlikely)<br>• 2<br>• 3<br>• 4 (Very likely) |
| 22. In order to increase the likelihood of myself ordering PGx test in the future for the management of my patients' drug therapy, I would need: * | Please rank the following statements with 1 being most useful and 9 being least useful<br>• Better knowledge of genetics<br>• Better knowledge of pharmacology<br>• Insurance coverage for patients |

| | |
|---|---|
| | • More traction within Singapore medical community<br>• Support from my working institution<br>• Available live assistance from an expert in genetics<br>• Better knowledge on medicinal chemistry (pharmacokinetics and pharmacodynamics)<br>• Stronger evidence that pharmacogenetics improves clinical outcomes<br>• Clearer legal regulations regarding PGx testing |
| 23. What do you think is missing from the current recruitment and on-boarding process? | • [Open-ended response] |
| 24. If you have more comments, please feel free to write them down below. | • [Open-ended response] |
